# Genome-wide meta-analysis of CSF biomarkers in Alzheimer’s disease and Parkinson’s disease cohorts

**DOI:** 10.1101/2023.06.13.23291354

**Authors:** Michael Ta, Cornelis Blauwendraat, Tarek Antar, Hampton L Leonard, Andrew B. Singleton, Mike A. Nalls, Hirotaka Iwaki, Alzheimer’s Disease Neuroimaging Initiative (ADNI), the Fox Investigation for New Discovery of Biomarkers

## Abstract

**Background:** Amyloid beta (Aβ), phosphorylated tau (p-tau), and total tau (t-tau) in cerebrospinal fluid are established biomarkers for Alzheimer’s disease (AD). In other neurodegenerative diseases, such as Parkinson’s disease (PD), these biomarkers have also been found to be altered, and the molecular mechanisms responsible for these alterations are still under investigation. Moreover, the interplay between these mechanisms and the diverse underlying disease states remains to be elucidated.

**Objectives:** To investigate genetic contributions to the AD biomarkers and assess the commonality and heterogeneity of the associations per underlying disease status.

**Methods:** We conducted GWAS for the AD biomarkers on subjects from the Parkinson’s Progression Markers Initiative (PPMI), the Fox Investigation for New Discovery of Biomarkers (BioFIND), and the Alzheimer’s Disease Neuroimaging Initiative (ADNI) and meta-analyzed with the largest AD GWAS.[7] We tested heterogeneity of associations of interest between different disease statuses (AD, PD, and control).

**Results:** We observed three GWAS signals: the *APOE* locus for Aβ, the 3q28 locus between *GEMC1* and *OSTN* for p-tau and t-tau, and the 7p22 locus (top hit: rs60871478, an intronic variant for *DNAAF5*, also known as *HEATR2*) for p-tau. The 7p22 locus is novel and co-localized with the brain *DNAAF5* expression. While no heterogeneity from underlying disease status was observed for the above GWAS signals, some disease risk loci suggested disease specific associations with these biomarkers.

**Conclusions:** Our study identified a novel association at the intronic region of *DNAAF5* associated with increased levels of p-tau across all diseases. We also observed some disease specific genetic associations with these biomarkers.

## Introduction

Alzheimer’s disease (AD) poses a significant social burden globally.[1, 2] Cerebrospinal fluid (CSF) levels for amyloid beta (Aβ), phosphorylated Tau (p-tau), and total Tau (t-tau) are established AD biomarkers integrated into the NIA-AA (National Institute on Aging – Alzheimer’s Association) research framework for Alzheimer’s disease.[16] The pathological significance of these biomarkers has been studied, and genome-wide association studies (GWAS) for these biomarkers have identified several GWAS loci associated with AD risk and progression.[6, 7, 9, 19, 26] CSF Aβ levels have been shown to be lower in AD cases whereas levels of CSF p-tau are elevated compared to normal subjects. Interestingly, these biomarkers were also reported to be altered in Parkinson’s disease (PD).[13, 17, 37] Previous studies that directly explored the relationship between these CSF biomarkers and PD showed a decrease in levels of both CSF Aβ and p-tau in cases versus control subjects. However, the genetic background of these observations across neurodegenerative diseases have not been well investigated.

In this study, we conducted GWAS on the AD biomarkers of participants from PD focused studies: the Parkinson’s Progression Markers Initiative (PPMI)[24] and the Fox Investigation for New Discovery of Biomarkers (BioFIND). We combined these results with the largest GWAS conducted on mixed cohorts for AD and controls.[7] We stratified the analysis on the recruitment study arms of each cohort and assessed the overall genetic contributions across healthy controls and case subjects for AD and PD groups irrespective of clinical phenotype. In addition, we investigated disease specific genetic contributions through the genetic heterogeneity between individuals with PD, AD, and healthy volunteers using the above PD studies as well as the Alzheimer’s Disease Neuroimaging Initiative (ADNI). We also assessed the genetic associations with the biomarker changes overtime when longitudinal data were available.

## Methods

### Participants

PPMI is an ongoing longitudinal observational study with multiple study arms. The current analyses included data from participants with early-stage idiopathic PD but had not yet received medication for PD at enrollment (PPMI_PD), healthy controls (PPMI_HC), those with scans without evidence of dopaminergic deficit but with Parkinsonism (PPMI_SWEDD), and those with prodromal symptoms such as hyposmia, REM sleep behavior disorder, and image confirmed dopaminergic deficit (PPMI_PRODROMAL). This study also included two genetically enriched study arms from PPMI where carriers of any high risk or causal variant for Parkinson’s disease (*LRRK2* G2019S, R1441C/G, *GBA1* N409S, L483P, 84GG, and *SNCA* A53T) were recruited. Both carriers with PD less than 7 years from diagnosis (PPMI_GENPD) and unaffected carriers or their 1st degree family members (PPMI_GENUN) were analyzed. BioFIND was a cross-sectional study with two study arms: PD cases in moderately advanced stages (BioFIND_PD) and healthy controls (BioFIND_HC). Both of these study arms were included in this study. The protocols for these studies can be obtained from the Michael J. Fox Foundation for Parkinson’s research (https://www.michaeljfox.org). We also included participants from the Alzheimer’s Disease Neuroimaging Initiative (ADNI, https://adni.loni.usc.edu). Based on their last diagnosis, these participants were stratified as either having dementia, (ADNI-Dementia), mild cognitive impairment (ADNI-MCI), or normal cognition (ADNI-NC). While clinical diagnosis of AD can change over time, 97% of the ADNI-Dementia were “probable” AD by NINCDS/ADRDA criteria according to the last record when available. Clinical data of the study participants such as disease status, age, sex, age at diagnosis were obtained from the study websites on December 12th, 2021. For a summary of the study design and data used across disease statuses, please refer to **Supplemental Figure 1**. Descriptive statistics for each study are available in **Supplementary Table 1**.

### CSF biomarkers

For ADNI and PPMI samples, CSF concentrations of Amyloid-β 1to42 (Aβ), total tau (t-tau), and phosphorylated tau at the threonine 181 position (p-tau) were measured using Elecsys electro-chemiluminescence immunoassays on the cobas e 601 analysis platform (Roche Diagnostics).[28] For BioFIND samples, these biomarkers were measured by INNO-BIA AlzBio3 immunoassay.[23] The detailed procedures and quality control process are summarized on the study websites. For GWAS analyses, biomarker values were log transformed and centered at zero to be compatible with existing summary statistics.

### Genetic data

We used the whole genome sequencing data (WGS) provided by the ADNI repository and the AMP-PD project.[15] The samples were sequenced (30x or more coverage) and underwent the GATK best practices workflow. Additional details regarding quality control are provided on the study websites. In this analysis we used PASS filtered variants and analyzed only the participants with European ancestry because of insufficient power to analyze non-European ancestry groups. The ancestry was confirmed by being within +/- 6SD of the first two principal components of the European samples (CEU and TSI) in HapMap3 panel.[12] We also excluded related individuals closer than 2nd degree relatives from the analysis.

### Summary statistics from the prior GWAS

We requested the summary statistics from the largest GWAS for Aβ, t-Tau and p-Tau from NIAGADS (https://www.niagads.org/) data repository (NG00055).[7] The GWAS included 3,146 individuals with and without dementia from nine different studies conducted at the Charles F. and Joanne Knight Alzheimer’s Disease Research Center (Knight ADRC), Saarland University in Homburg/Saar, Germany (HB), Mayo Clinic (MAYO), Skåne University Hospital, Sweden (SWEDEN), Perelman School of Medicine at the University of Pennsylvania (UPENN), and the University of Washington (UW) as well as Alzheimer’s Disease Neuroimaging Initiative (ADNI1 and ADNI2) and Predictors of Cognitive Decline Among Normal Individuals (BIOCARD). The CSF biomarkers were log-transformed and centered per study followed by a single-stage association test adjusted for age, sex, measurement platform, and the first two principal components.

### Analysis

Cohort-strata-level GWAS for the AD biomarkers were conducted. For the BioFIND study, we fit a linear regression model for additive allele effect using the cross-sectional data. For the other studies where longitudinal data were available, we used the GALLOP algorithm to approximate the linear mixed effects model for both the additive allele effect (cross-sectional associations) and the additive *allele x time* interaction (longitudinal associations). This algorithm provides equivalent solutions to a linear mixed effects model in a computationally efficient way.[29] In both models, we adjusted for age, sex, and the first two principal components (PC1-PC2). For the GALLOP model, we further adjusted for time from the baseline measurement, interactions between time and PCs (PC1-PC2), and a random intercept and random slope for each individual. Our primary analysis was to meta-analyze the cross-sectional results with the previously reported summary statistics from the largest CSF AD biomarker GWAS[7] to identify the across-disease genetic contributions for these biomarkers. We also meta-analyzed the longitudinal associations to see if there are any genome-wide significant loci associated with the biomarker change over time.

In the meta-analysis, all of the variants with a minor allele count less than 5 or minor allele frequency <1% among the individual studies, not reported in more than 2 study arms, or failed for heterogeneity assessment (p-value for the test of heterogeneity < 0.05 or I2 > 80%) were removed for from the “overall” genetic assessment for the biomarkers. For novel genome-wide significant loci, we further assessed co-localization with brain eQTL (expression quantitative trait loci)[3] and blood eQTL[33] using LocusCompare.[21]

For those SNPs reported in the previous study by Deming et al, we assessed if there was any evidence of heterogeneity between different disease states. We meta-analyzed GWAS from PPMI-PD, PPMI-GENPD, BioFIND-PD to compose “PD” GWAS summary statistics. Similarly, we meta-analyzed ADNI-CN, BioFIND-HC, and PPMI-HC and PPMI-GENUN to generate “HC” GWAS results. For AD, we used the ADNI-Dementia GWAS summary statistics. The heterogeneity between these disease specific GWAS results were assessed using their I^2^ statistics.

Finally, we assessed if disease specific or nonspecific genetic associations with these CSF biomarkers exist for known risk loci associated with either PD or AD.[4, 22] For this targeted analysis, the significance level was set at the false positive rate (q-value) of 0.05 adjusting for the number of loci to be tested.

All the statistical analyses and drawings were executed using Plink version 2.0 alpha,[25] R version 3.6 and Python version 3.8. Meta-analyses were conducted using METAL software with an inverse variance weighted method with the genomic control correction applied.[36] The analysis scripts are available at https://github.com/NIH-CARD/biomarker_longGWAS. The data were obtained from ADNI and AMP-PD.

## Results

We used data from 61 GWAS in the main meta-analyses, including previously published results: 34 GWAS for cross-sectional components and 27 GWAS for longitudinal components. The genomic inflation factors of the GWAS were reasonable, with most around 1.0 except for three between 1.1-1.3 <u>(Supplemental Table 2)</u>. The inflation was accounted for in the meta-analysis phase by the genomic control function in METAL.

In the meta-analysis, we observed three GWAS signals in the cross-sectional component: the *APOE* locus for Aβ, the 3q28 locus between *GEMC1* and *OSTN* for p-tau and t-tau, and the 7p22 locus (top hit: rs60871478, an intron variant for *DNAAF5*, also known as *HEATR2*) for p-tau <u>(Table 1</u>, Figure 1, <u>Supplemental Figure 2 and 3</u>). No genome-wide signals were identified in the longitudinal components <u>(Supplemental Figure 5)</u>. The rs769449 variant on the APOE locus was identified to be significant for all three CSF biomarkers. For rs35055419, a significant association was seen for p-tau and t-tau. Both of these variants were previously reported by Cruchaga et. al.

**Figure 1.**
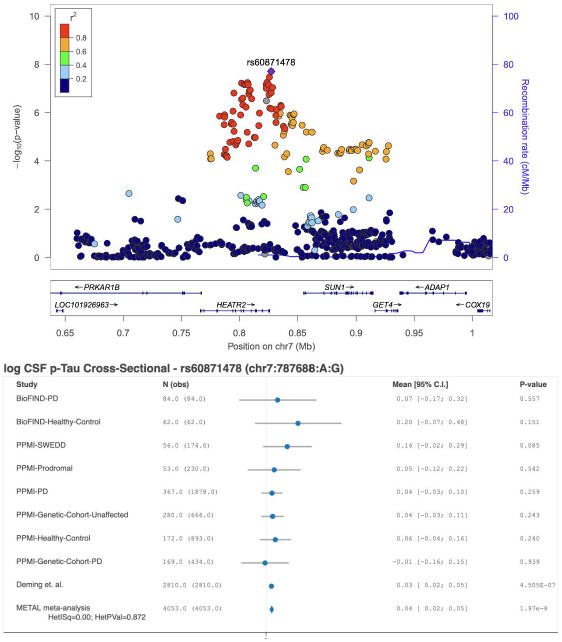
rs60871478 locus for log CSF p-tau and forest plots.

The chromosome 7 locus in the *DNAAF5* gene region was not previously reported and showed genome-wide significant P-value for p-tau P=1.97E-8 and a sub genome-wide significant P-value for t-tau P=5.82E-7. When exploring the potential causal gene in this region we identified eQTLs for *DNAAF5* that were well-colocalized with the GWAS signals in both the brain and blood (Supplemental Figure 4). No other genes in this locus showed any colocalization between the QTL and the GWAS signal.

Disease specific genetic associations with CSF biomarkers was assessed on the significant loci reported by Deming et al. between AD and PD related dementia. A stratified meta-analysis by disease state suggested heterogeneity was present for one of the non-replicated associations between rs12961169 (*CTDP1*) and p-tau. The ADNI-Dementia group showed a negative association (p=0.00182) that was not observed in the PD and the HC groups (Supplemental Figure 6 and 7). No evidence of heterogeneity for the other non-replicated associations was seen. One reported locus at 1p32.3 for Aβ (rs185031519, very rare) was not identified in the current analysis.

Likewise, for known AD and PD risk associated loci on CSF biomarkers, the disease stratified meta-analysis was used to identify the genetic similarities and differences across the dementias and control groups. The *APOE* e4 tagging allele was associated with lower CSF Aβ Regardless of the disease status (Table 2). AD risk increasing allele, rs6586028_T (*TSPAN14*) was associated with the increasingly lower CSF Ab over time in the PD group but not in the ADNI-Dementia and the HC groups, although there was not enough evidence suggesting heterogeneity among these results. The PD risk increasing allele, rs7134559_C (12q13.11) was associated with lower p-tau in a disease status non-specific way.

## Discussion

In this study, we conducted GWAS on CSF levels of three known AD biomarkers. We used data from PD and AD studies and assessed multiple diseases and stages of progression. We replicated two genetic loci from the previous largest AD biomarker study[7] (*APOE* and *GEMC1*) and identified a new locus at 7p22 that reached genome-wide significance in association with p-tau. *APOE* was previously shown to have significant associations with CSF biomarkers and is a genetic risk factor for late-onset sporadic AD[5]. In PD, carriers of the *APOE*-ε4 allele were found to have both quicker cognitive decline compared to non-carriers and an increased risk of progression to dementia.[30] Prior studies have found rs9877502 on the 3q28 locus between *GEMC1* and *OSTN* to be associated with higher CSF tau levels and identified a risk variant (rs1316356) for AD that is in linkage disequilibrium with this SNP.[6, 7] Additionally, *GEMC1* has been recently reported to be a key molecule in multiciliated cell differentiation.[20] In the brain, these cells are involved in maintaining homeostasis and neurogenesis. The new 7p22 locus, rs60871478, was associated with increased levels of CSF p-tau regardless of the disease status and colocalized well with *DNAAF5* (also known as *HEATR2*). When exploring the potential causal gene in this region we identified that there was good colocalization between the GWAS signal and blood and brain eQTL data for *DNAAF5* and no correlation was seen with other genes in the locus.

This gene encodes the protein Dynein Axonemal Assembly Factor 5 and is essential for the pre-assembly or stability of axonemal dynein. A missense mutation in *DNAAF5* was identified in a whole-exome sequencing study of a family with primary ciliary dyskinesia, a rare autosomal recessive disease that presents with neonatal respiratory distress, sinopulmonary disease, otitis media, male infertility, and left-right laterality defects. The affected individuals showed a malfunction in airway epithelial cells.[11] The gene is expressed ubiquitously across all tissues however the link between p-tau and the gene is unclear.

To assess the clinical consequence of the variant, we conducted ad-hoc analyses testing associations of this locus with age at onset and MMSE in the ADNI cohort <u>(Supplemental Table 3)</u>. The results suggest the loci is associated with MMSE scores in the ADNI-Dementia group (−1.07±0.48, p-val: 0.025). A similar association was shown in the ADNI-CN group with a smaller magnitude of effect (−0.34±0.17, p-val: 0.021). This variant may play a role in cognitive decline related to increased tau pathology with underlying AD pathology, but the sample size is not large enough to adjust for multiple testing and further evaluation with a larger cohort would be required. In addition, biological evaluation such as cellular or animal Alzheimer models may provide more information regarding progression and dementia risk associated with the locus.

Six previously reported cross-sectional associations with CSF biomarkers in AD were not replicated in this study. Of these, rs12961169 (*CTDP1*) showed high heterogeneity across diseases. For the others, it may in part be due to differences in the study designs. The previous study was focused on identifying AD related loci using the biomarkers as endophenotypes. So, they conducted one-stage GWAS without adjusting for the disease status.

Interestingly, the PD risk increasing loci rs7134559 (*SCAF11*) was associated with the lower CSF p-tau. A recent study reported that these biomarkers were indeed lower in PD when compared with HC in contrast to the generally higher CSF p-tau in AD.[13] The reason for the difference in these biomarker profiles between the two diseases is unknown, but there are many reports suggesting the influence of AD pathology to the PD pathology or vice versa.[14, 27, 35] The current observation may be associated with some interaction between the two disease mechanisms.

Longitudinal changes are smaller compared to baseline differences in biomarker levels between disease states. Reported heterogeneity on CSF biomarker trajectories has been observed in AD risk allele carriers prior to and after the onset of dementia symptoms.[8] The need for a stratified analysis by dementia progression in addition to larger cohort sizes might improve detection of longitudinal genetic contributions.

By integrating data from PD studies, we were able to expand the knowledge of genetic-biomarker relationships that were mainly derived from AD studies previously. For some SNPs, we could differentiate disease specific and non-specific genetic associations. Admittingly, the sizes of this study, especially in regards to the disease specific GWAS, were still small. Additional data is needed for more effective analyses in particular large datasets from diverse ancestries with longitudinal measures available. Nevertheless, we believe that the current approach would be useful to investigate underlying AD mechanisms modified by different disease status. Another limitation of this study is the potential misdiagnosis of AD and PD because the clinical diagnosis is not always accurate. In particular, misdiagnosis would affect the heterogeneity assessment. Access to additional biomarkers and increasing study sizes are both important to overcome this problem.

We also like to note that this study may have not fully accounted for the complex relationships between Aβ, p-tau, and t-tau. These biomarkers are recognized as endophenotypes, and they are highly sensitive and specific in differentiating AD and controls.[5, 10] However, the clinical significance of these biomarkers is not equal. First, they are thought to represent different pathological processes related to AD: low CSF Aβ for aggregation of Aβ, high CSF pTau for neurofibrillary tangle (NFT) formation, and high CSF tTau for neurodegeneration.[32] The timing of deviation from normal is also different, as the decrease of Aβ is observed earlier than the increase of CSF p-Tau and t-Tau.[10] Additionally, the increase of tTau and pTau in the early stage of AD may be associated with faster progression of disease.[31, 34] Moreover, there is a study that reported the level of p-Tau, supposedly reflecting tangle pathology, was more closely associated with amyloid PET than with tau PET.[18] To further investigate these complex relationships between the biomarkers, GWAS on various stratifications, such as disease status, other biomarker status and imaging status, should provide important information to untangle these relationships.

In conclusion, we analyzed the CSF AD biomarker from the AD and the PD studies. We identified three associations across disease status including one novel genome-wide significant locus and also observed some associations suggesting the disease specific modifications of these biomarkers at known risk loci.

## Data Availability

All data produced in the present study are available upon reasonable request to the authors

## Acknowledgement

The research team would like to thank the participants and individuals involved in the organization and collection of study materials.

This work was supported by the Center for Alzheimer’s and Related Dementias, within the Intramural Research Program of the National Institute on Aging and the National Institute of Neurological Disorders and Stroke, National Institutes of Health, Department of Health and Human Services. Project numbers Z01-AG000957-19 and Z01-AG000534-03. This work utilized the computational resources of the NIH HPC Biowulf cluster. (http://hpc.nih.gov).

Data collection and sharing for this project was funded by the Alzheimer’s Disease Neuroimaging Initiative (ADNI) (National Institutes of Health Grant U01 AG024904) and DOD ADNI (Department of Defense award number W81XWH-12-2-0012). ADNI is funded by the National Institute on Aging, the National Institute of Biomedical Imaging and Bioengineering, and through generous contributions from the following: AbbVie, Alzheimer’s Association; Alzheimer’s Drug Discovery Foundation; Araclon Biotech; BioClinica, Inc.; Biogen; Bristol-Myers Squibb Company; CereSpir, Inc.; Cogstate; Eisai Inc.; Elan Pharmaceuticals, Inc.; Eli Lilly and Company; EuroImmun; F. Hoffmann-La Roche Ltd and its affiliated company Genentech, Inc.; Fujirebio; GE Healthcare; IXICO Ltd.;Janssen Alzheimer Immunotherapy Research & Development, LLC.; Johnson & Johnson Pharmaceutical Research & Development LLC.; Lumosity; Lundbeck; Merck & Co., Inc.;Meso Scale Diagnostics, LLC.; NeuroRx Research; Neurotrack Technologies; Novartis Pharmaceuticals Corporation; Pfizer Inc.; Piramal Imaging; Servier; Takeda Pharmaceutical Company; and Transition Therapeutics. The Canadian Institutes of Health Research is providing funds to support ADNI clinical sites in Canada. Private sector contributions are facilitated by the Foundation for the National Institutes of Health (www.fnih.org). The grantee organization is the Northern California Institute for Research and Education, and the study is coordinated by the Alzheimer’s Therapeutic Research Institute at the University of Southern California. ADNI data are disseminated by the Laboratory for Neuro Imaging at the University of Southern California.

Data used in the preparation of this article were obtained from the Parkinson’s Progression Markers Initiative (PPMI) database (www.ppmi-info.org/access-data-specimens/download-data). For up-to-date information on the study, visit www.ppmi-info.org. PPMI – a public-private partnership – is funded by the Michael J. Fox Foundation for Parkinson’s Research and funding partners, including: 4D Pharma; AbbVie Inc.; AcureX Therapeutics; Allergan; Amathus Therapeutics; Aligning Science Across Parkinson’s (ASAP); Avid Radiopharmaceuticals; Bial Biotech; Biogen; BioLegend; Bristol Myers Squibb; Calico Life Sciences LLC; Celgene Corporation; DaCapo Brainscience; Denali Therapeutics; The Edmond J. Safra Foundation; Eli Lilly and Company; GE Healthcare; GlaxoSmithKline; Golub Capital; Handl Therapeutics; Insitro; Janssen Pharmaceuticals; Lundbeck; Merck & Co., Inc.; Meso Scale Diagnostics, LLC; Neurocrine Biosciences; Pfizer Inc.; Piramal Imaging; Prevail Therapeutics; F. Hoffmann La Roche Ltd and its affiliated company Genentech Inc.; Sanofi Genzyme; Servier; Takeda Pharmaceutical Company; Teva Neuroscience, Inc.; UCB; Vanqua Bio; Verily Life Sciences; Voyager Therapeutics, Inc.; and Yumanity Therapeutics, Inc.

BioFIND is funded by The Michael J. Fox Foundation for Parkinson’s Research and the National Institute Neurological Disorders and Stroke.

