## Supplemental Figure for "Genome-wide meta-analysis of CSF biomarkers in Alzheimer’s disease and Parkinson’s disease cohorts"

### Supplemental Figure 1. Analysis overview

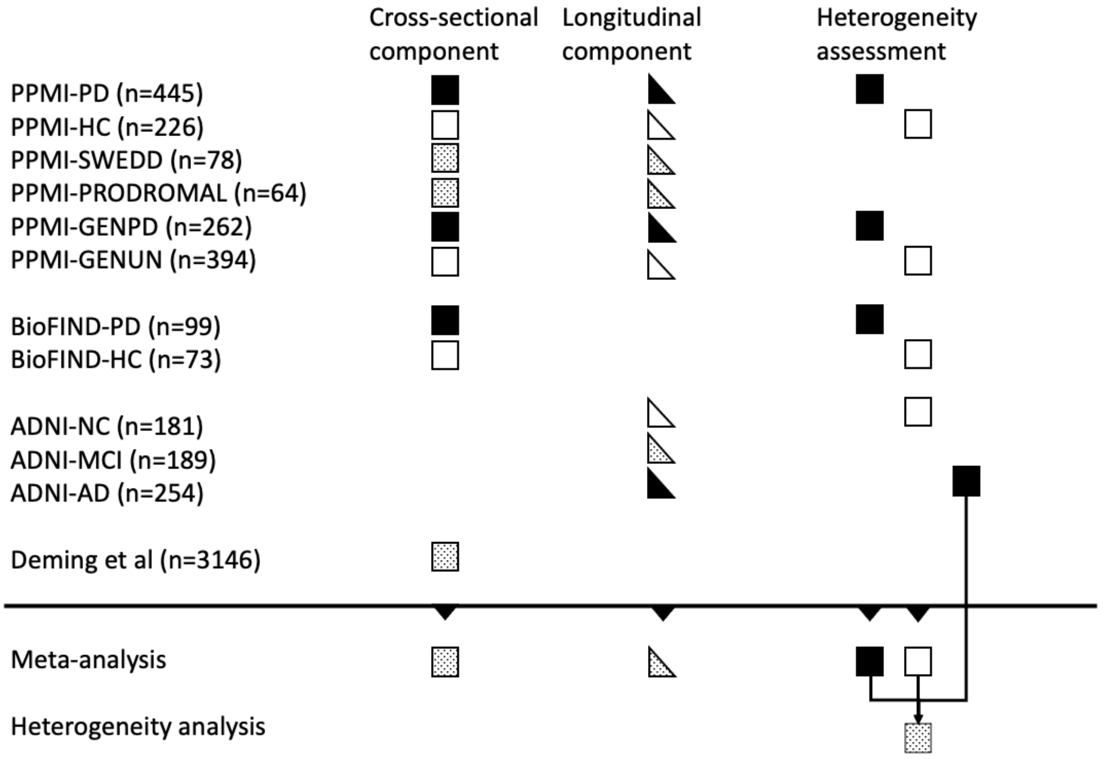


Individual arms were analyzed separately and then meta-analyzed. Squares represent the cross-sectional component analysis results while the triangles represent the longitudinal component (change over time) analysis results. The disease groups (either with Parkinson's disease or Alzheimer's disease) were filled with black while non-disease groups were unfilled. Dotted fills indicate the results from those with mixed/non-typical diseases. As shown in the reverse triangles we meta-analyzed cross-sectional component GWAS results and longitudinal component GWAS results retrospectively. In the heterogeneous analyses, we first meta-analyzed GWAS results per disease status for PD and HCs and then compared them.

PPMI, Parkinson's Progression Marker Initiative; BioFIND, the Fox Investigation for New Discovery of Biomarkers; ADNI, the Alzheimer's Disease Neuroimaging Initiative; PPMI-PD, PD patients in the PPMI study; PPMI-HC, healthy controls in the PPMI study; PPMI-SWEDD,  those with scans without evidence of dopaminergic deficit in the PPMI study; PPMI-PRODROMAL, those with prodromal symptoms such as hyposmia, REM sleep behavior disorder, and image confirmed dopaminergic deficit in the PPMI study; PPMI_GENPD, GBA/LRRK2/SNCA variant carriers of PD with Parkinson's disease; PPMI-GENUN, the unaffected GBA/LRRK2/SNCA variant carriers without PD; BioFIND-PD, cases in moderately advanced stages (BioFIND_PD) in the BioFIND study; BioFIND-HC, healthy controls in the BioFIND study. ADNI-AD, those with dementia, (ADNI-AD) in the ADNI study; ADNI-MCI, those with mild cognitive impairment; and ADNI-NC, those with normal cognition (ADNI-NC).

### Supplemental Figure 2. GWAS results for cross-sectional component

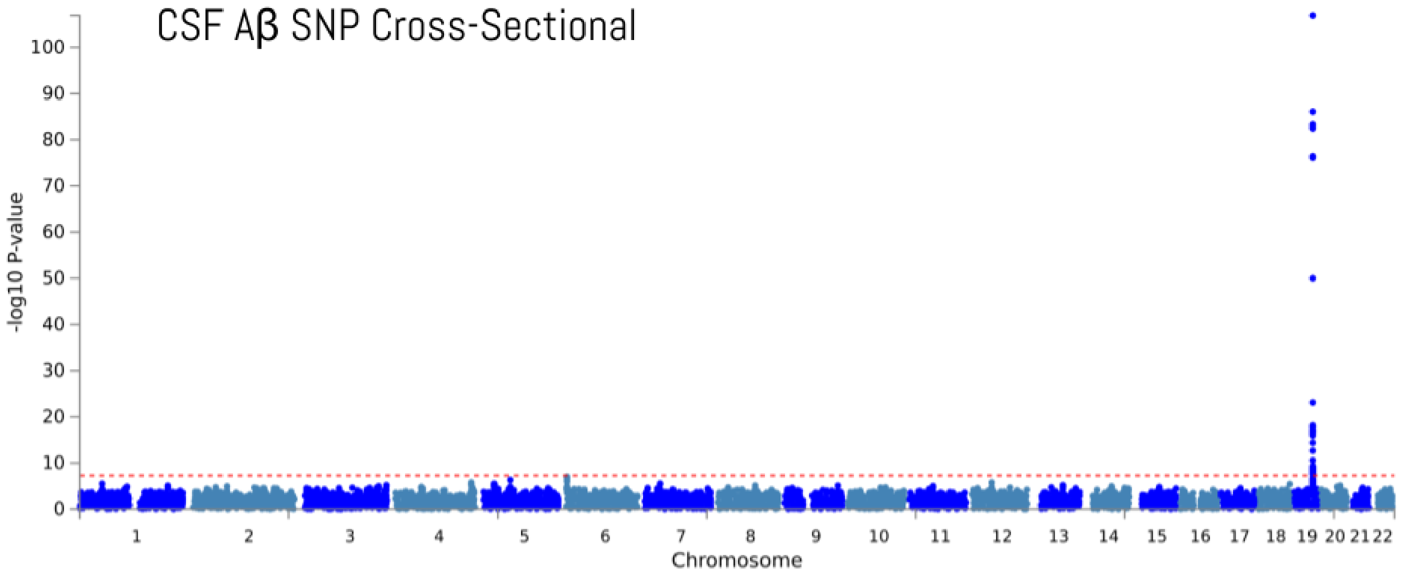


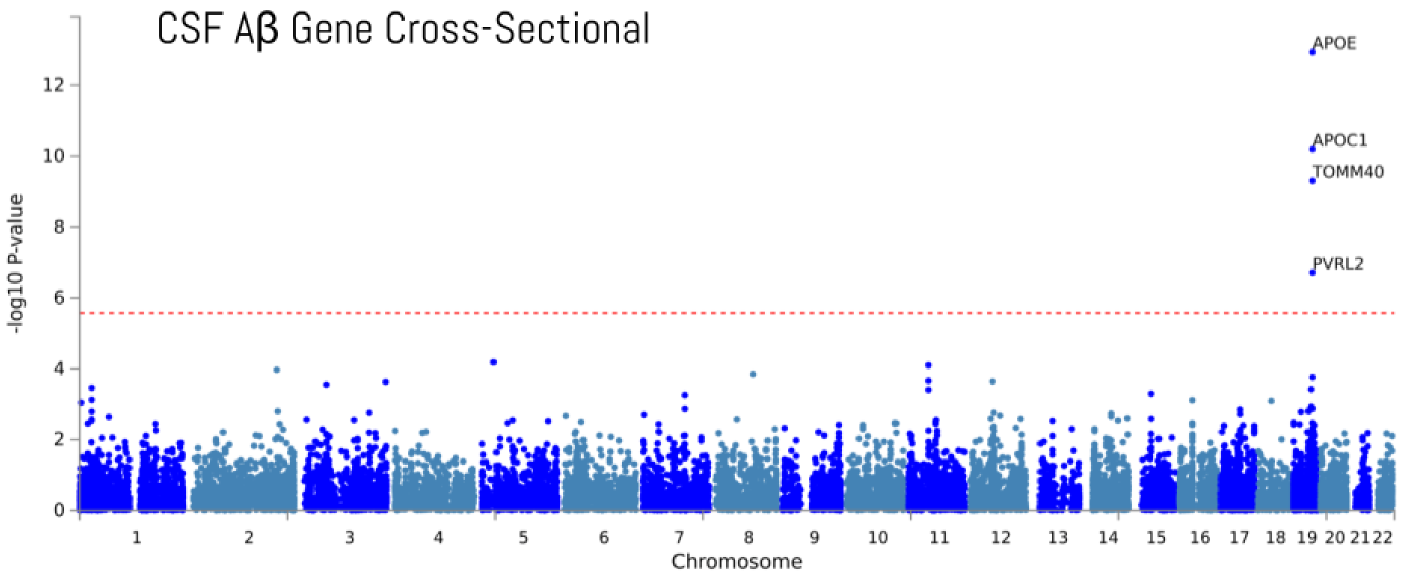

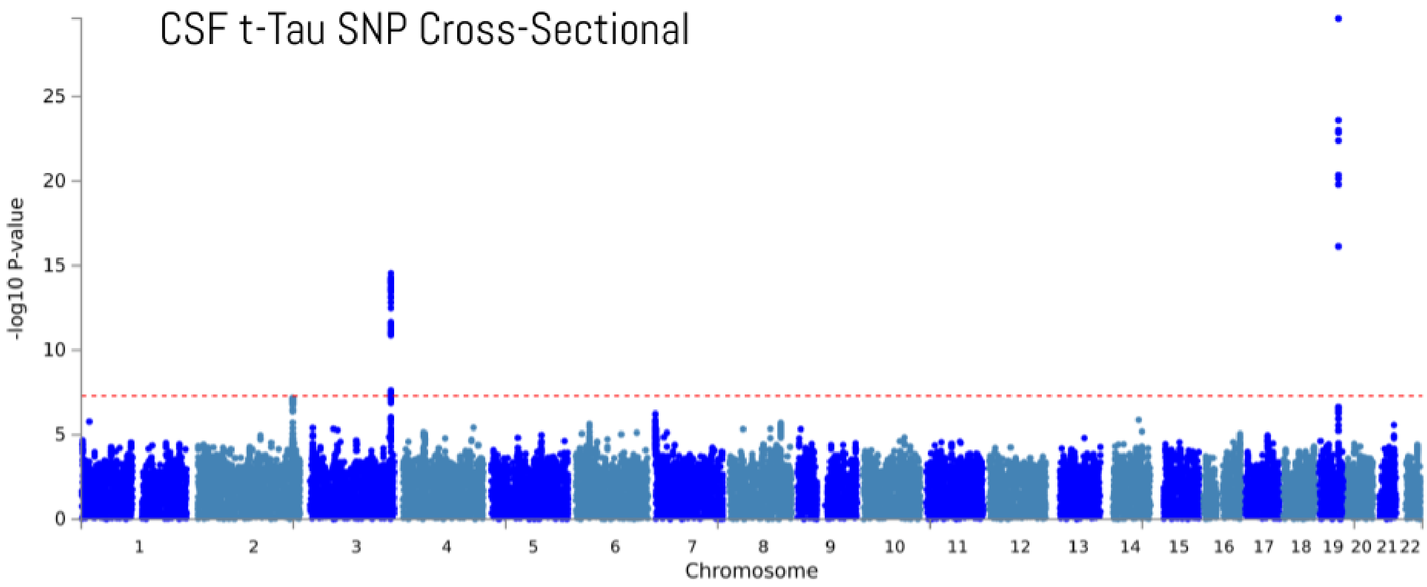
CSF t-Tau SNP Cross-Sectional
-log10 P-value
0
5
10
15
20
25
1 2 3 4 5 6 7 8 9 10 11 12 13 14 15 16 17 18 19 20 21 22
Chromosome

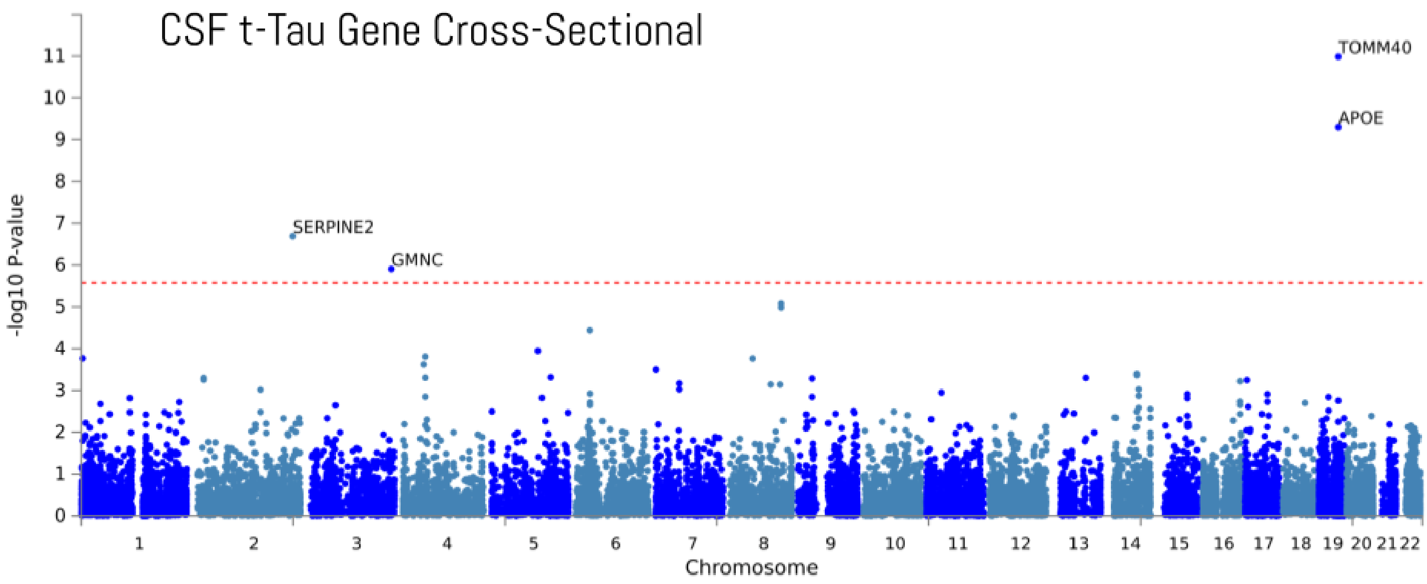
CSF t-Tau Gene Cross-Sectional
TOMM40
APOE
SERPINE2
GMNC
-log10 P-value
0
1
2
3
4
5
6
7
8
9
10
11
1 2 3 4 5 6 7 8 9 10 11 12 13 14 15 16 17 18 19 20 21 22
Chromosome

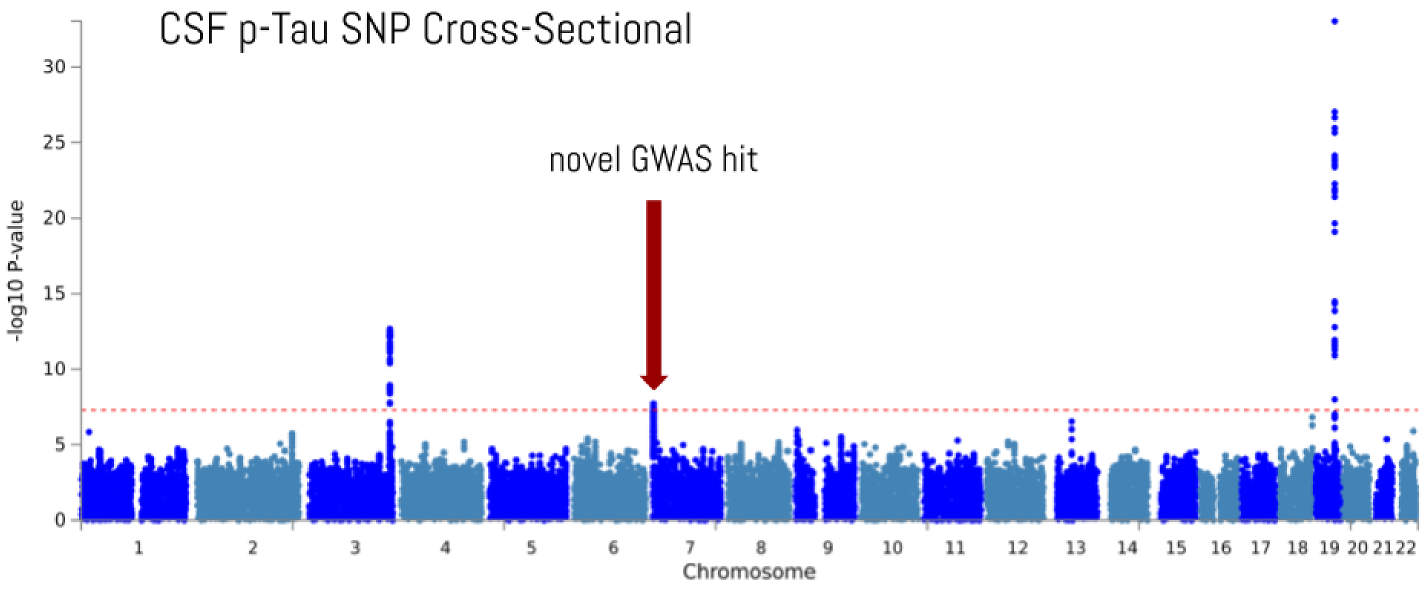
CSF p-Tau SNP Cross-Sectional
novel GWAS hit
-log10 P-value
Chromosome

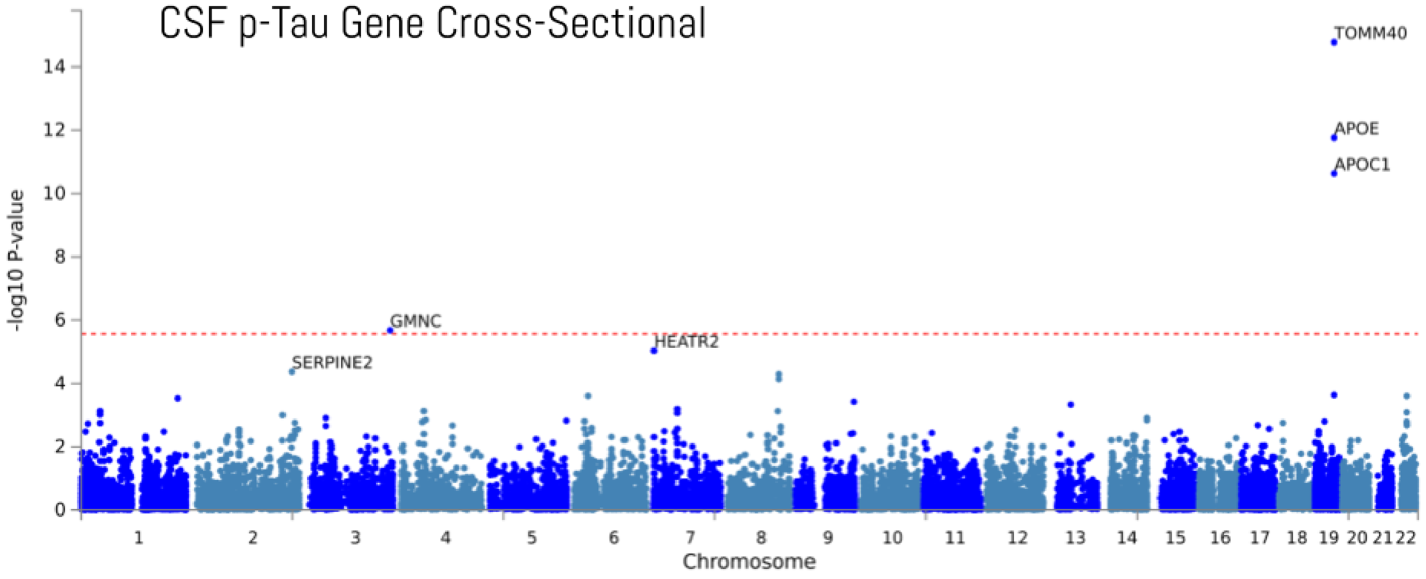
CSF p-Tau Gene Cross-Sectional
TOMM40
APOE
APOC1
GMNC
HEATR2
SERPINE2
-log10 P-value
Chromosome

### Supplemental Figure 3. Forest plot for the GWAS loci - rs35055419 and rs769449

#### log CSF Aβ Cross-Sectional - rs769449 (chr19:44906745:G:A)

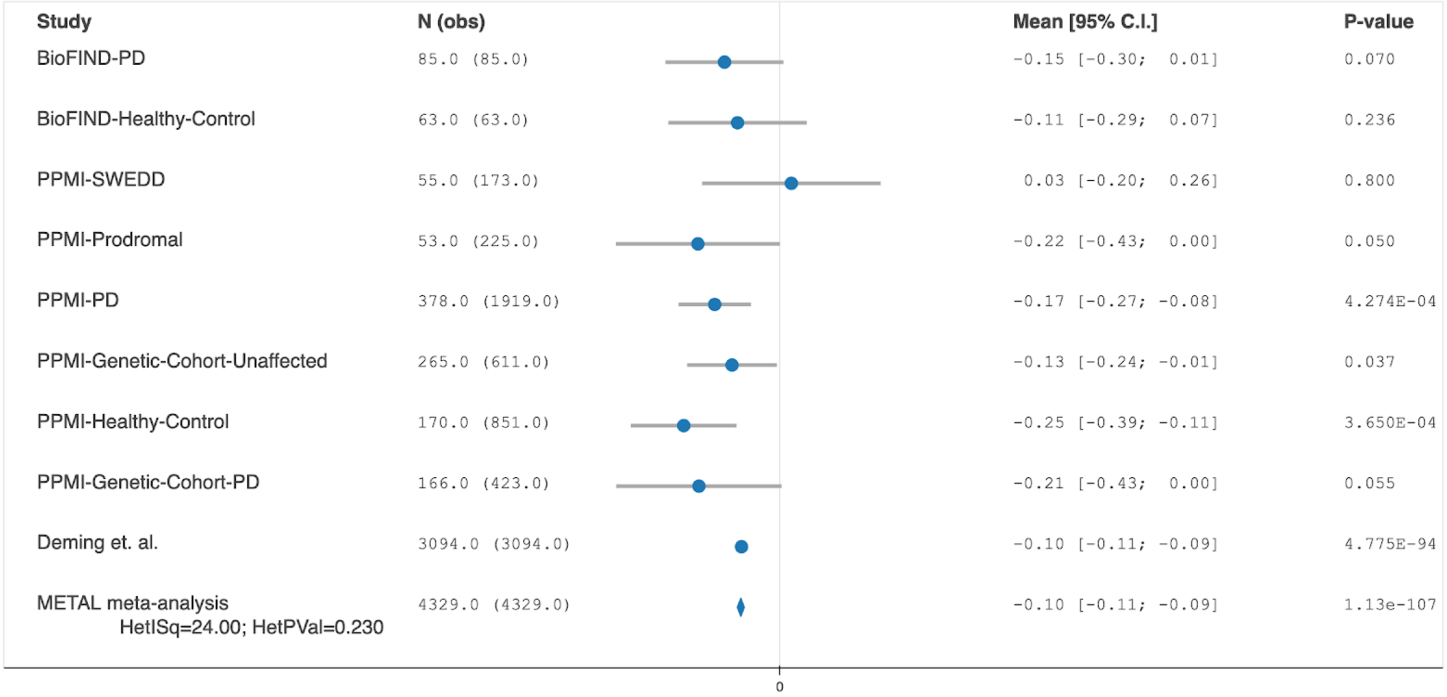


#### log CSF p-Tau Cross-Sectional - rs35055419 (chr3:190945768:T:C)

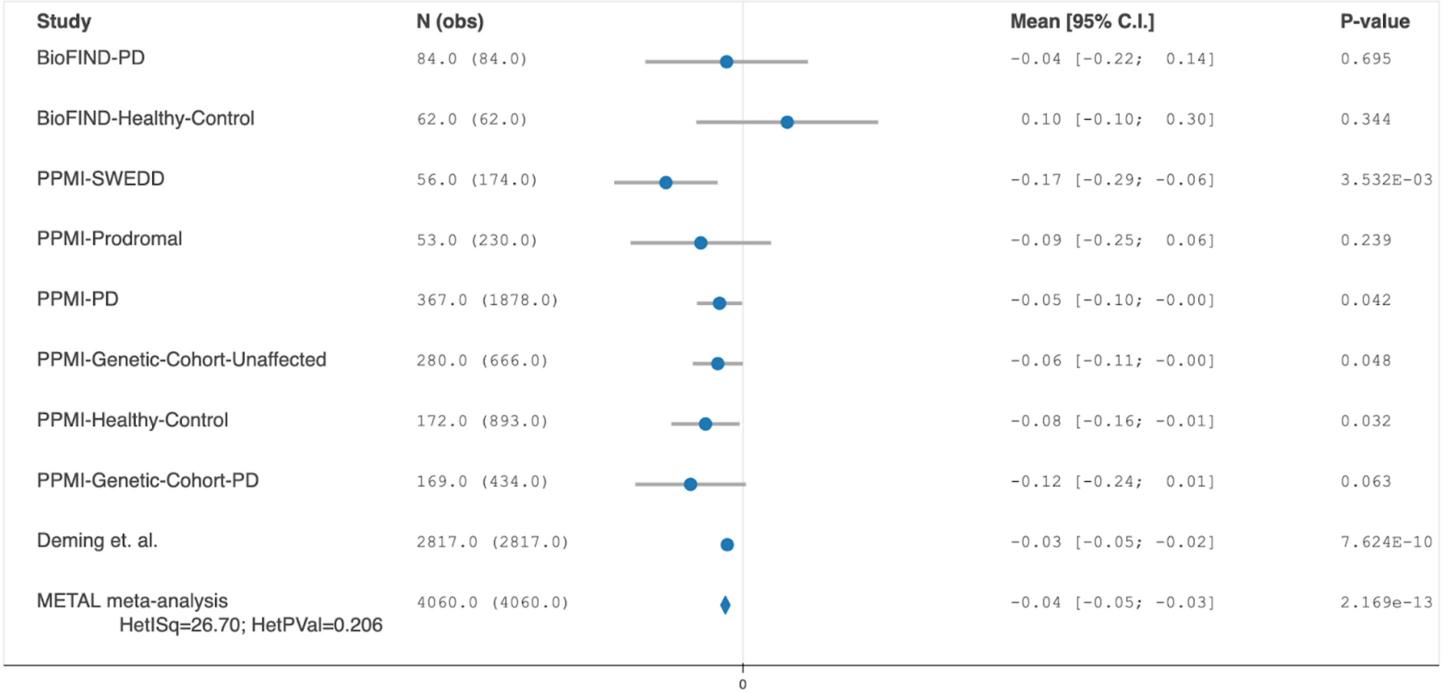

#### log CSF t-Tau Cross-Sectional - rs35055419 (chr3:190945768:T:C)

| Study | N (obs) | Mean [95% C.I.] | P-value |
|---|---|---|---|
| BioFIND-PD | 85.0 (85.0) | -0.02 [-0.15; 0.12] | 0.823 |
| BioFIND-Healthy-Control | 62.0 (62.0) | 0.02 [-0.11; 0.14] | 0.796 |
| PPMI-SWEDD | 57.0 (178.0) | -0.16 [-0.27; -0.05] | 4.390E-03 |
| PPMI-Prodromal | 54.0 (239.0) | -0.07 [-0.22; 0.08] | 0.376 |
| PPMI-PD | 378.0 (2014.0) | -0.05 [-0.10; -0.00] | 0.040 |
| PPMI-Genetic-Cohort-Unaffected | 283.0 (676.0) | -0.06 [-0.11; -0.00] | 0.047 |
| PPMI-Healthy-Control | 175.0 (942.0) | -0.10 [-0.17; -0.02] | 8.316E-03 |
| PPMI-Genetic-Cohort-PD | 173.0 (456.0) | -0.10 [-0.22; 0.03] | 0.129 |
| Deming et. al. | 3091.0 (3091.0) | -0.04 [-0.05; -0.03] | 3.071E-11 |
| METAL meta-analysis<br>HetISq=4.50; HetPVal=0.397 | 4358.0 (4358.0) | -0.04 [-0.05; -0.03] | 2.748e-15 |

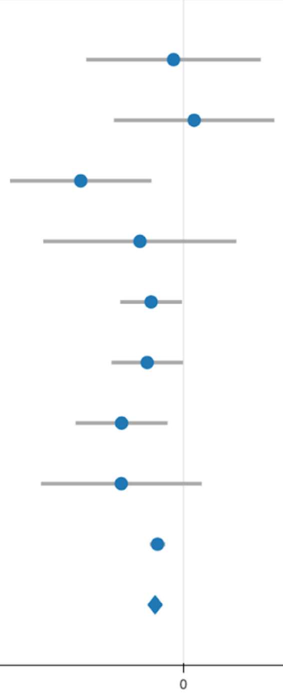

### Supplemental Figure 4. Co-localization plots for the chromosome 7 and

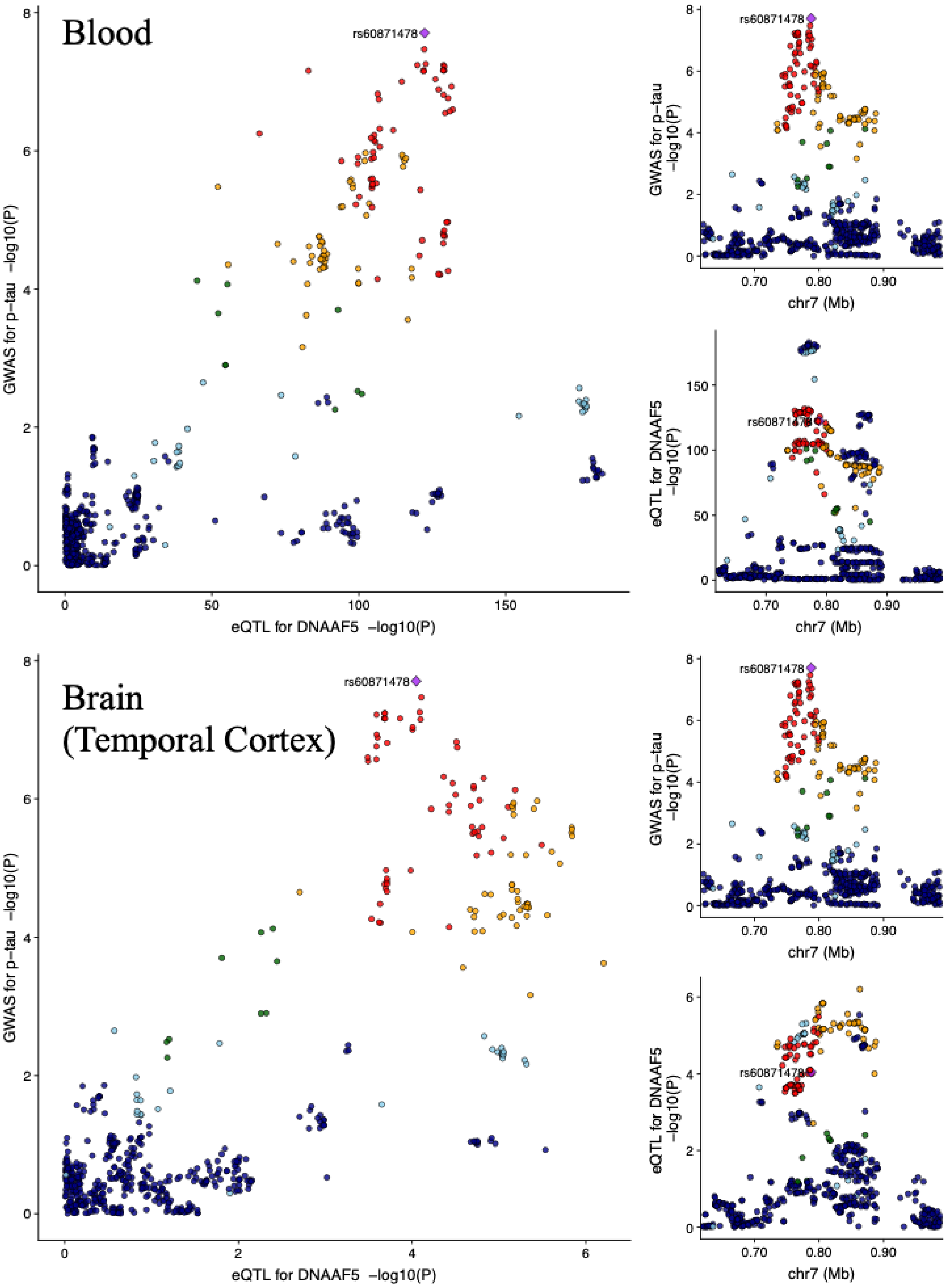

Supplemental Figure 5. GWAS results for longitudinal component

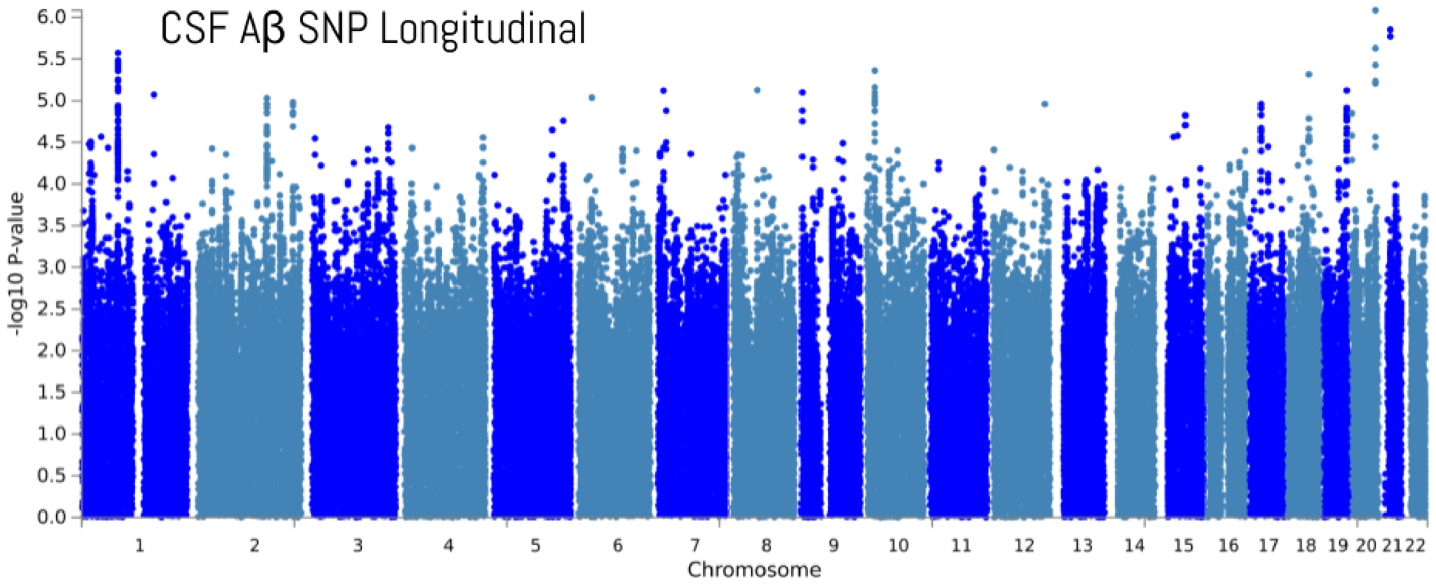


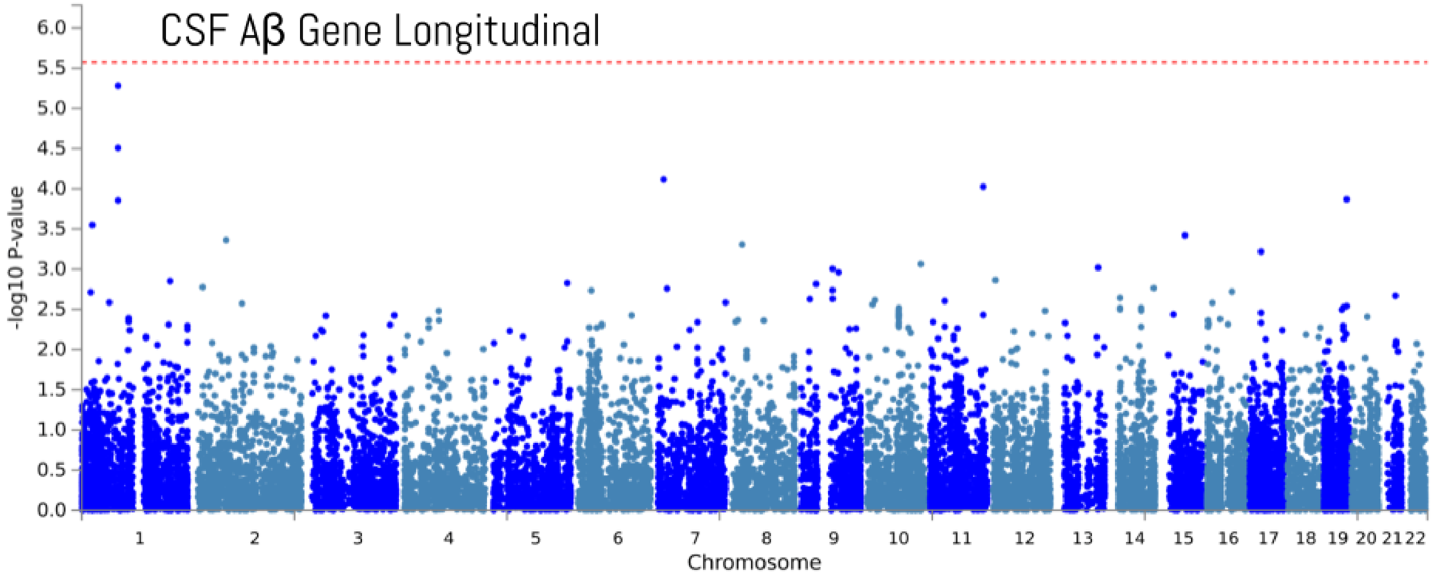

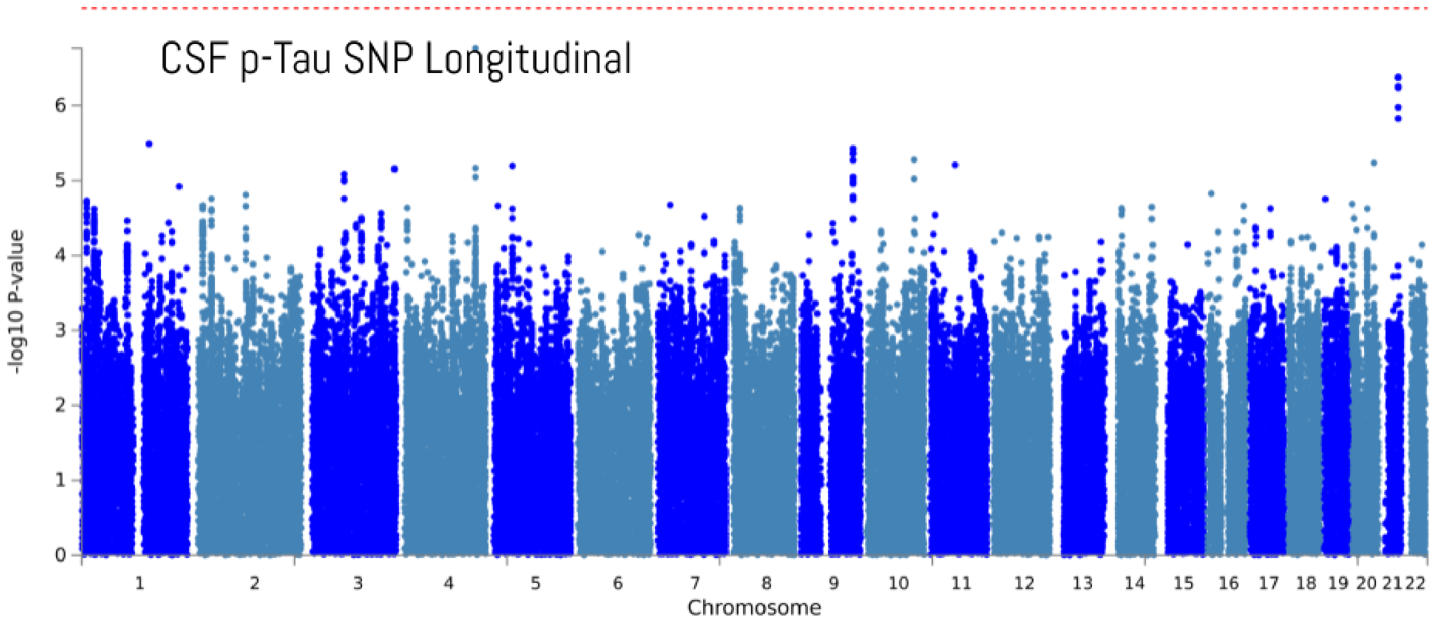
CSF p-Tau SNP Longitudinal
-log10 P-value
Chromosome
0
1
2
3
4
5
6
1 2 3 4 5 6 7 8 9 10 11 12 13 14 15 16 17 18 19 20 21 22

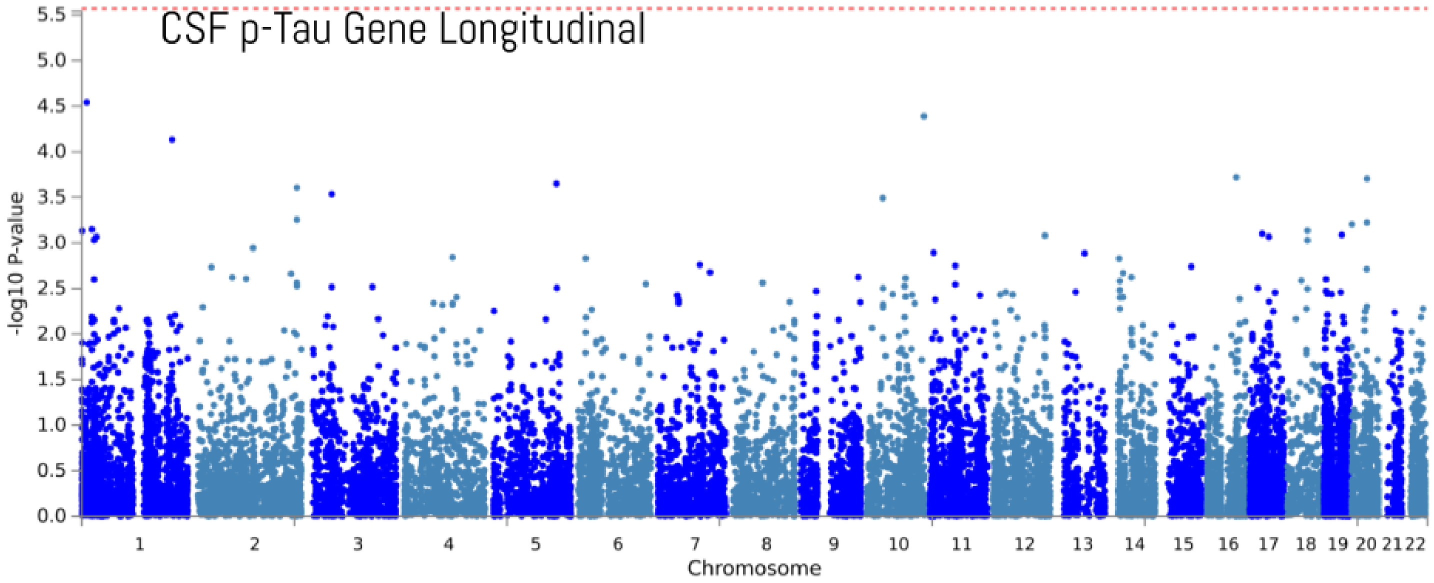
CSF p-Tau Gene Longitudinal
-log10 P-value
Chromosome
0.0
0.5
1.0
1.5
2.0
2.5
3.0
3.5
4.0
4.5
5.0
5.5
1 2 3 4 5 6 7 8 9 10 11 12 13 14 15 16 17 18 19 20 21 22

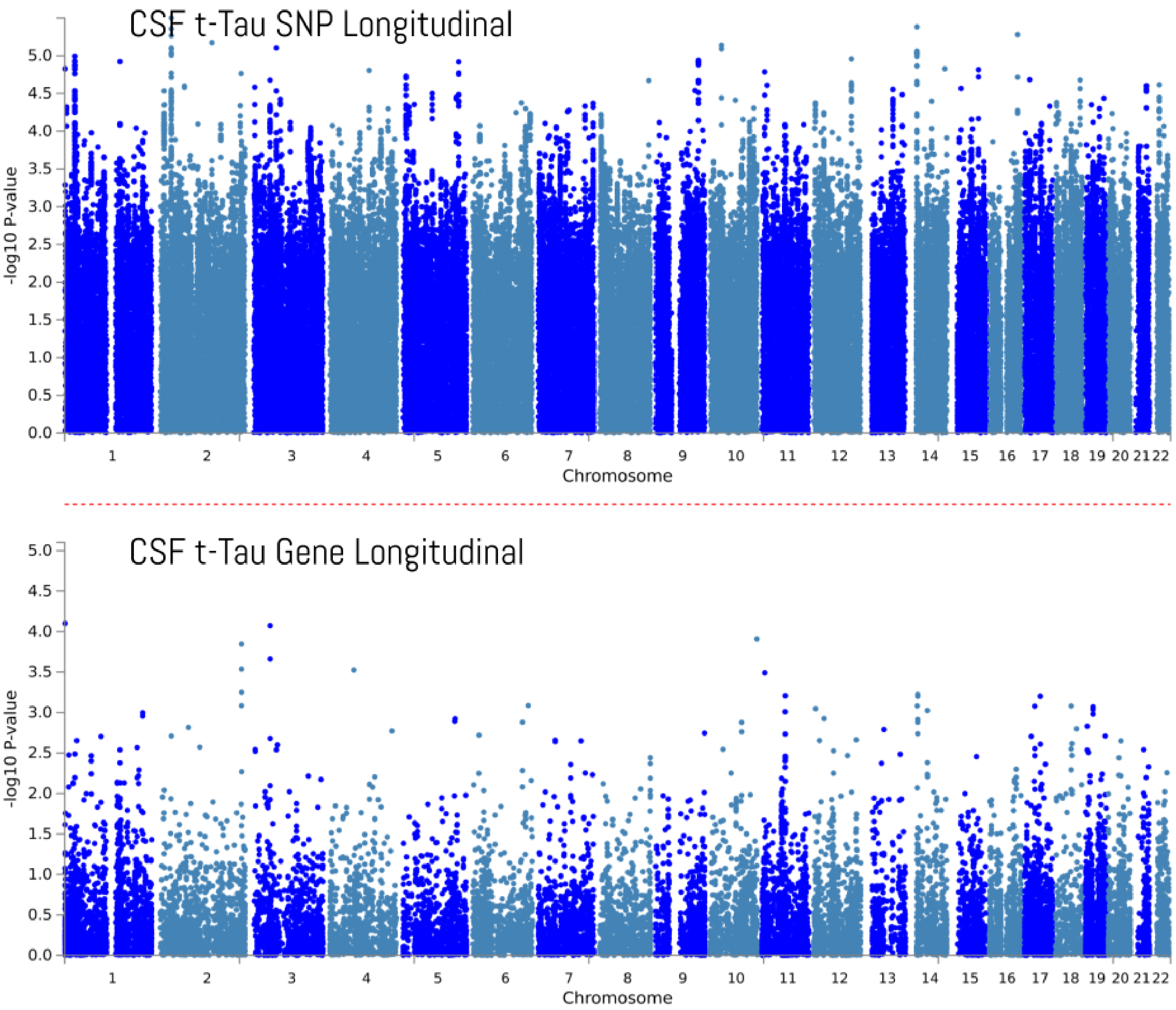
CSF t-Tau SNP Longitudinal
-log10 P-value
0.0 0.5 1.0 1.5 2.0 2.5 3.0 3.5 4.0 4.5 5.0
1 2 3 4 5 6 7 8 9 10 11 12 13 14 15 16 17 18 19 20 21 22
Chromosome
CSF t-Tau Gene Longitudinal
-log10 P-value
0.0 0.5 1.0 1.5 2.0 2.5 3.0 3.5 4.0 4.5 5.0
1 2 3 4 5 6 7 8 9 10 11 12 13 14 15 16 17 18 19 20 21 22
Chromosome

Supplemental Figure 6. UpSet plots per disease status for the association previously reported by Deming et al.

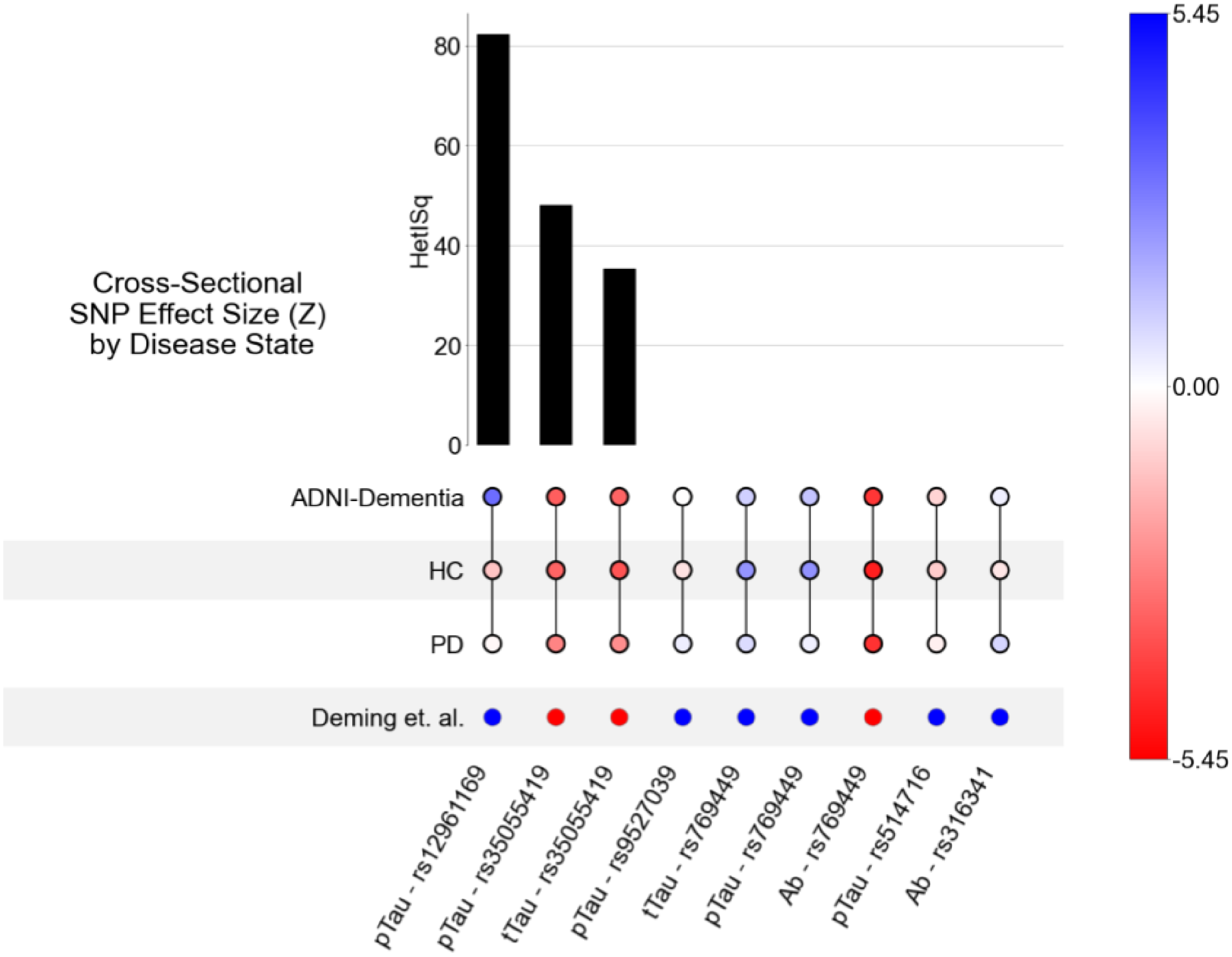


One reported loci at 1p32.3 for Aβ (rs185031519) was not identified in the current dataset.

### Supplemental Figure 7. Forest plot for SNP - rs12961169

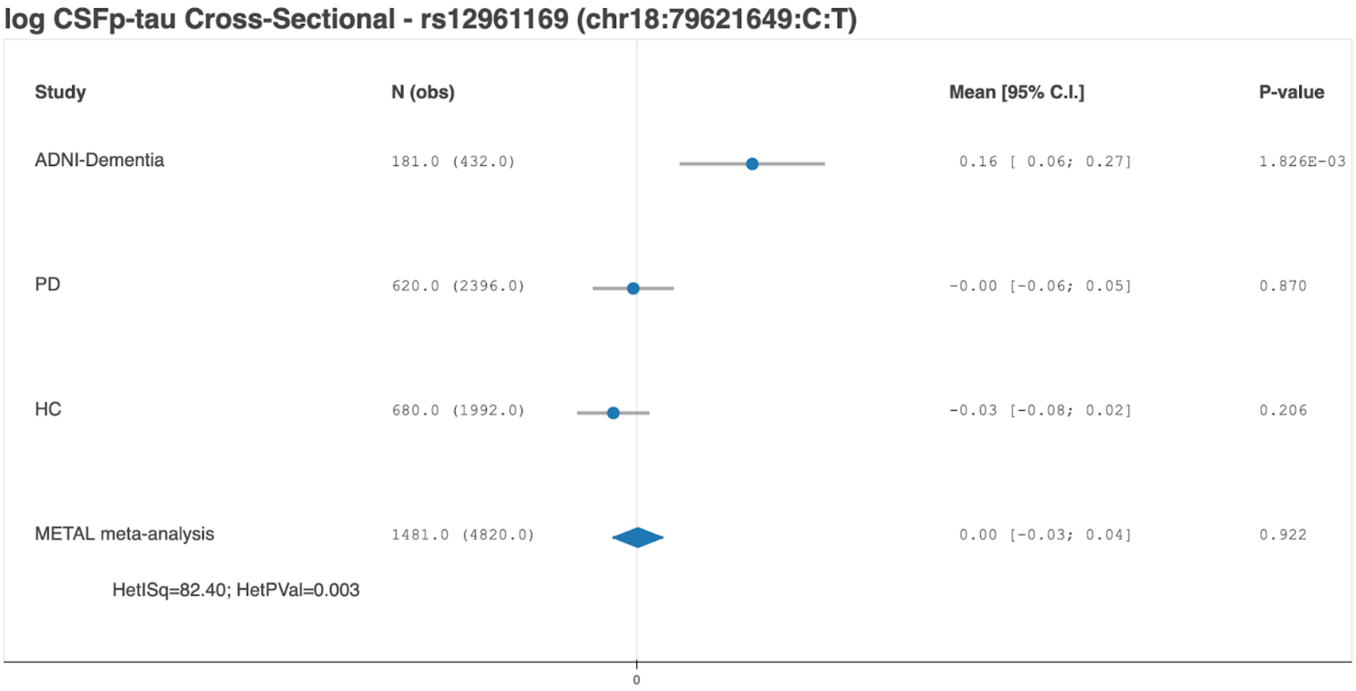
